# Effectiveness and duration of a second COVID-19 vaccine booster

**DOI:** 10.1101/2022.10.03.22280660

**Authors:** Alejandro Jara, Cristobal Cuadrado, Eduardo A. Undurraga, Christian García, Manuel Nájera, María Paz Bertoglia, Verónica Vergara, Jorge Fernández, Heriberto Garcia, Rafael Araos

## Abstract

Using a prospective national cohort of 3.75 million individuals aged 20 or older, we evaluated the effectiveness against COVID-19 related ICU admissions and death of mRNA-based second vaccine boosters for four different three-dose background regimes: BNT162b2 primary series plus a homologous booster, and CoronaVac primary series plus an mRNA booster, a homologous booster, and a ChAdOx-1 booster. We estimated the vaccine effectiveness weekly from February 14 to August 15, 2022, by estimating hazard ratios of immunization over non-vaccination, accounting for relevant confounders. The overall adjusted effectiveness of a second mRNA booster shot was 88.2% (95%CI, 86.2-89.9) and 90.5% (95%CI 89.4-91.4) against ICU admissions and death, respectively. Vaccine effectiveness showed a mild decrease for all regimens and outcomes, probably associated with the introduction of BA.4 and BA.5 Omicron sub-lineages and immunity waning. The duration of effectiveness suggests that no additional boosters are needed six months following a second booster shot.

## Main text

The COVID-19 pandemic has caused more than 615 million cases and 6.5 million deaths reported globally as of September 2022.^1^ COVID-19 vaccines have been essential to decrease the burden of disease and reduce restrictions associated with the pandemic response. A robust body of evidence showed that the primary series of several COVID-19 vaccines had high efficacy and effectiveness against symptomatic COVID-19 and severe illness in the first months.^2,3^ However, immunity waning and the emergence of new, highly transmissible SARS-CoV-2 lineages with increased immune evasion, lead many countries to roll out booster shots four to six months following the primary series.^4,5^ Research showed that recently administered homologous or heterologous COVID-19 boosters restored waning protection against symptomatic infection and severe illness.^6-8^

The emergence and spread of the B.1.1.529 Omicron variant of SARS-CoV-2, and its sub-lineages, caused an unprecedented global surge in COVID-19 cases.^1^ As happened with the primary series of COVID-19 vaccines, research showed substantial reductions in protection against symptomatic COVID-19.^9-12^ Early in 2022, several countries began rolling out second boosters (a fourth dose for most vaccines), four to six months following the first booster dose, and international organizations have recommended its use for at-risk populations.^13-17^ Policymakers need evidence on real-world effectiveness to guide vaccination policies. However, there is limited evidence for the effectiveness of second boosters, and it primarily refers to mRNA vaccines in Israel, Canada, and the USA among populations at higher risk (e.g., older adults and immunocompromised persons).^12,15,17^ Furthermore, there is no evidence of a potential waning of protection against COVID-19 for second boosters, or the effectiveness against severe disease of an additional booster dose for individuals who received their primary series based on inactivated vaccines, which constitute a considerable proportion of vaccinated individuals in low- and middle-income countries.^18^

Using a large prospective national observational cohort in Chile, we evaluated the effectiveness of mRNA-based second vaccine boosters for individuals with four different three-dose background regimes: (i) BNT162b2 primary series plus a homologous booster (3mRNA), (ii) CoronaVac primary series plus mRNA booster (CC+mRNA), (iii) CoronaVac primary series plus homologous booster (CCC), and (iv) CoronaVac primary series plus ChAdOx-1 booster (CCA). We estimated vaccine effectiveness weekly, from February 14, 2022, to August 15, 2022, against admission to an intensive care unit (ICU) and death (U07.1) associated with laboratory-confirmed SARS-CoV-2 infection. We estimated the overall vaccine effectiveness of an mRNA second booster, regardless of the three-dose background regimen, and for each group, using survival regression models to estimate hazard ratios of immunization (>13 days after the second dose) over non-vaccination, accounting for time-varying vaccination exposure and clinical, demographic, and socioeconomic confounders at baseline.

The Chilean Ministry of Health launched the second booster campaign on February 14, 2022, based on a standard dose of Pfizer-BioNTech’s BNT162b2 or Moderna’s mRNA-1273 vaccines, prioritizing the elderly and immunocompromised individuals. By August 29, 2022, more than 10.6 million individuals had received an additional booster dose, representing 71.7% of the target population. Vaccination rollout and the health system in Chile have been described elsewhere.^19^

Our study cohort included adults aged ≥20 years and affiliated with the Fondo Nacional de Salud (FONASA), the public national healthcare system, who completed a CoronaVac or BNT162b2’s two-dose primary series at least 120 days before the beginning of the follow-up on August 11, 2021, when the first-booster campaign was launched, and unvaccinated individuals. We excluded individuals with probable or confirmed COVID-19 according to reverse-transcription polymerase-chain-reaction assay for SARS-CoV-2 or antigen test reported before February 15, 2022 (Extended Fig.1). Our final cohort included 3,754,785 adults. Of these, 2,623,802 (69.9) received a second booster shot of BNT162b2 or mRNA-1273 vaccine between February 14, 2022, and August 18, 2022, and 757,726 (20.2%) had not been vaccinated by the end of the follow-up. Vaccination rollout was organized through a publicly available schedule and was free of charge (Extended Fig.2). Notably, there was a high level of SARS-CoV-2 circulation during the rollout, in which the predominant Omicron sub-lineages were BA.2.1.12, BA.4, and BA.5 (Extended Fig.3). Cohort characteristics are described in Supplementary Material Tables 1-5. The incidence of COVID-19 and the vaccination status at the end of the follow-up differed significantly (*P*<0.001) by participant’s sex, age group, comorbidities, nationality, region of residence, and income.

**Table 1.** Effectiveness against ICU admissions and death of mRNA-based second vaccine boosters in adults aged 20 years and older, February 14, 2022, through August 15, 2022*.

| Immunization status | Persons at risk | Person-days | Cases |  | Vaccine effectiveness (95% CI) |
| --- | --- | --- | --- | --- | --- |
|  |  |  | No. | Age and sex adjusted incidence (100 thousand person-days) | End of follow-up (August 15, 2022) |
| Admitted to ICU |  |  |  |  |  |
| Unvaccinated | 749,856 | 137,442,303 | 388 | 0.293<br>(0.264-0.323) | -<br>- |
| Overall | 2,602,987 | 300,876,487 | 296 | 0.054<br>(0.043-0.065) | 88.2<br>(86.2-89.9) |
| 3mRNA | 236,895 | 23,626,638 | 16 | 0.069<br>(0.028-0.110) | 74.2<br>(54.8-85.2) |
| CC+mRNA | 840,136 | 89,423,892 | 76 | 0.064<br>(0.048-0.080) | 77.0<br>(69.7-83.0) |
| CCC | 137,002 | 19,206,580 | 37 | 0.053<br>(0.032-0.074) | 74.2<br>(54.8-85.2) |
| CCA | 1,393,752 | 203,137,349 | 210 | 0.037<br>(0.030-0.044) | 86.3<br>(83.2-88.8) |
| Confirmed deaths |  |  |  |  |  |
| Unvaccinated | 749,188 | 137,223,419 | 1,115 | 0.684<br>(0.642-0.725) | -<br>- |
| Overall | 2,603,731 | 300,919,202 | 552 | 0.081<br>(0.071-0.092) | 90.5<br>(89.4-91.4) |
| 3mRNA | 236,930 | 23,629,947 | 9 | 0.146<br>(0.010-0.282) | 87.7<br>(69.7-83.0) |
| CC+mRNA | 840,425 | 89,441,770 | 169 | 0.133<br>(0.112-0.154) | 81.0<br>(76.8-84.0) |
| CCC | 137,122 | 19,214,510 | 98 | 0.115<br>(0.092-0.138) | 79.3<br>(73.8-83.7) |
| CCA | 1,394,118 | 203,161,303 | 372 | 0.059<br>(0.052-0.066) | 90.8<br>(89.4-92.0) |

\* \*COVID-19 denotes coronavirus disease 2019. The Ministry of Health launched a COVID-19 vaccine first booster campaign on August 11, 2021, and a second booster campaign on February 14, 2022. The table shows the estimated vaccine effectiveness of mRNA-based second vaccine boosters for individuals with four different three-dose background regimes: i) BNT162b2 primary series plus a homologous booster (3mRNA), a CoronaVac primary series plus (ii) mRNA booster (CC+mRNA), (iii) homologous booster (CCC), or (iv) ChAdOx-1 booster (CCA), compared to unvaccinated individuals. Estimates were adjusted for time-varying vaccination exposure and clinical, demographic, and socioeconomic confounders at baseline (Supplementary Material Tables 1-5).

The overall age- and sex-standardized incidence of COVID-19-related ICU admissions and deaths among unvaccinated individuals was 0.29 per 100,000 person-days (95% confidence interval CI 0.26-0.32) and 0.68 (95% 0.64-0.75), respectively. In contrast, the adjusted incidence for ICU admissions and deaths for participants with a second booster dose was 0.05 per 100,000 person-days (95%CI 0.04-0.07) and 0.08 per 100,000 person-days (95%CI 0.07-0.09) (Table 1). At the end of follow-up, the overall adjusted vaccine effectiveness of a second booster to prevent ICU admission and death was 88.2% (95%CI 86.2-89.9) and 90.5% (95%CI 89.4-91.4), respectively. These estimates represent a moderate but significant reduction in vaccine effectiveness against ICU admissions and death from a maximum of 96.8% (95%CI 86.8-99.2) and death of 96.7% (95%CI 93.0-98.3), respectively (Fig.1 and Supplementary Material Tables 6-7).

**Figure 1.**
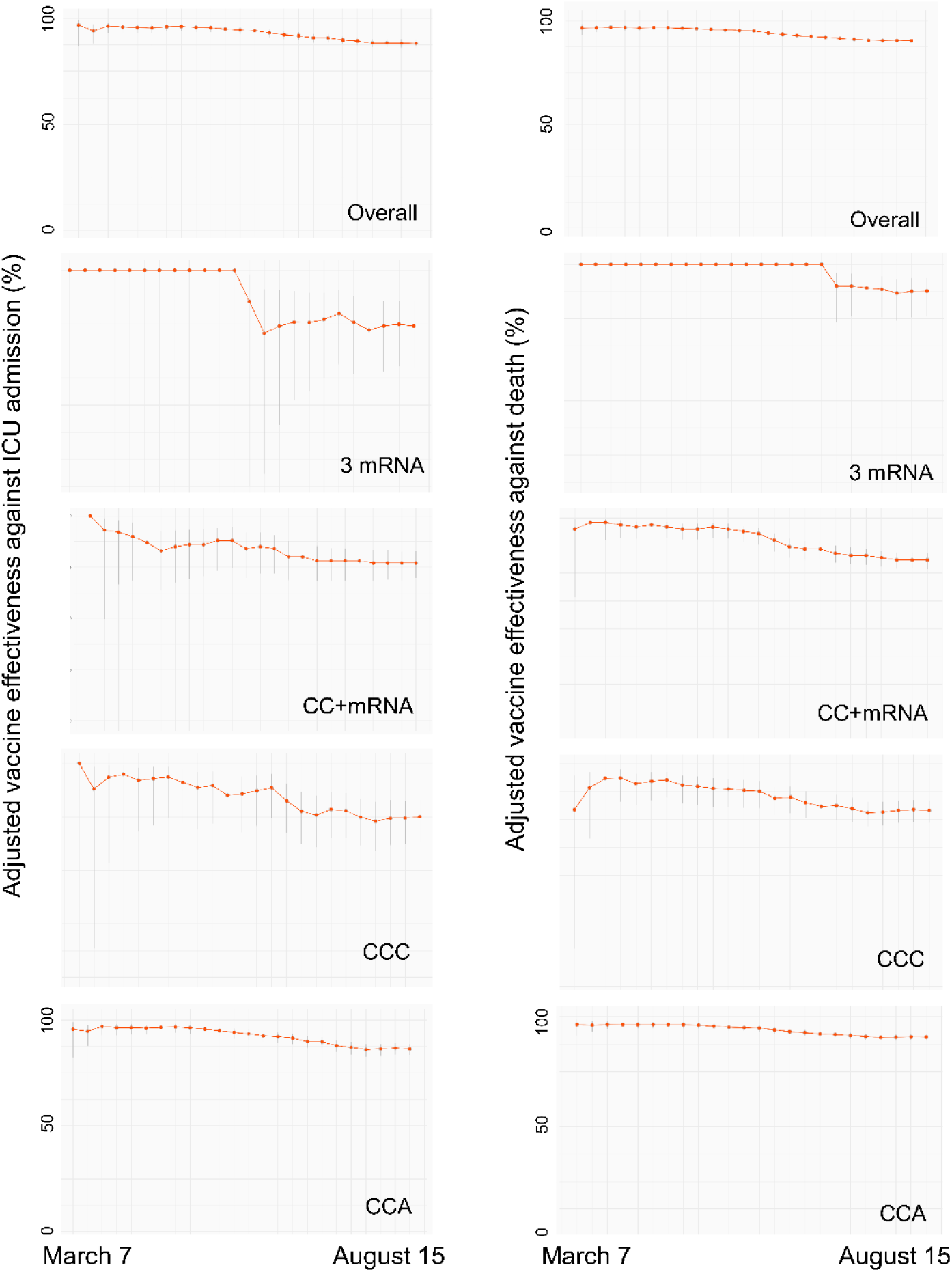
Overall vaccine effectiveness against COVID-19-related ICU admissions and death of mRNA-based second vaccine boosters for each week, March 7 through August 15, 2022. 3mRNA denotes individuals who received a BNT162b2 primary series plus a homologous first booster dose. CC+mRNA, CCC, and CCA denote individuals with a primary series of CoronaVac plus a first mRNA, homologous, and a ChAdOx-1 first booster dose, respectively.

The scheme-specific adjusted vaccine effectiveness of an additional booster dose against COVID-19 related ICU admission at the end of follow-up was 74.1% (95% CI 54.8-85.2), 75.0% (95%CI 63.4-83.0), 86.3% (95%CI 83.2-88.8), and 77.0 (95%CI 69.6-83.0) for 3mRNA, CCC, CC+mRNA, and CCA, respectively. The adjusted vaccine effectiveness against COVID-19 related deaths was 87.7% (95%CI 76.1-93.7), 79.3% (95%CI, 73.8-83.7), 81.0% (95%CI 76.8-84.0), and 90.8% (95%CI 89.4-92.0) for 3mRNA, CCC, CC+mRNA, and CCA, respectively. Vaccine effectiveness moderately decreased for all schemes and outcomes studied (Fig.1), although most confidence intervals overlapped, with some exceptions (Supplementary Material Tables 6-7): vaccine effectiveness against COVID-19 related ICU admissions significantly decreased for the CCA scheme, and protection against COVID-19 related deaths significantly decreased for CCC, CC+mRNA, and CCA (Fig.1 and Supplementary Material Tables 6-7).

Our cohort represents a population immunized with various schemes of COVID-19 vaccines. Ninety-one percent of the cohort received a primary series of CoronaVac, combined with a first booster based on a viral-vectored (53%) or an mRNA vaccine (37%). The remaining participants (9%) received a primary series of CoronaVac and a first homologous or BNT162b2 mRNA vaccine booster. Similar to the overall effect of a second mRNA booster dose, the scheme-specific vaccine effectiveness of these boosters remained high by the end of the follow-up, ranging from 74.2% to 86.3% effectiveness against COVID-19-related ICU admission and 79.3% to 90.8% against death. The CCA vaccination scheme plus the additional mRNA booster dose provided the highest protection against COVID-19-related deaths. Our study was not designed to demonstrate the superiority of any specific vaccination scheme. However, our results suggest that mix-and-match strategies may provide comparable protection against severe disease to homologous, mRNA-based regimens.

Our findings are consistent with previous studies on second booster doses. A recent summary of studies on mRNA vaccines,^17^ including eight studies from Israel, one from Canada,^12^ and one from the USA,^15^ all conducted during the Omicron outbreak, suggests that an additional mRNA booster enhances protection compared to three vaccine doses and unvaccinated individuals, particularly for severe outcomes. While not directly comparable, due to differences in study design, population characteristics, and outcome definitions, the studies in Canada^12^ and the USA^15^ estimate the vaccine effectiveness of an additional mRNA booster compared to unvaccinated individuals. The study in Canada, conducted among residents in long-term care facilities aged 60 years or older, found that the effectiveness of a fourth dose against severe outcomes was 87% (95%CI 82-90).^12^ In the USA, research was conducted among immunocompromised adults aged 50 years or older. Those results showed that the effectiveness of a fourth mRNA dose against hospitalization was 80% (95%CI 71-85). These studies have a relatively short follow-up time after the fourth dose, of two to ten weeks, and thus do not fully capture immunity waning.

At least two other studies have examined the effectiveness of additional booster doses. A preprint study in Thailand examined the effectiveness of an additional booster dose based on Oxford-AstraZeneca’s ChAdOx1 adenoviral vector vaccine among adults with various three-dose background regimes, including inactivated vaccines.^16^ The study found a vaccine effectiveness against symptomatic COVID-19 of 73% (95%CI 48-89) but did not examine its effectiveness against severe outcomes. Last, anecdotal evidence from two participants in a cohort of healthcare workers in the USA suggests that a second homologous mRNA booster may induce a substantial and durable neutralizing antibody response.^20^

Our study has some limitations. As an observational study, there is a risk of selection bias if the vaccinated and unvaccinated groups differ in a systematic way, such as risk aversion, which affects the probability of exposure to the treatment and the risk of SARS-CoV-2 infection. We adjusted our estimates with observable clinical, demographic, and socioeconomic confounders that could affect vaccination and the risk of infection, but there could be residual confounding. Second, our study design did not allow us to separate the effect of immunity waning, and the emergence of new SARS-CoV-2 lineages with increased immune evasion on our vaccine effectiveness estimates against severe disease for an additional mRNA booster dose. Results should be interpreted with caution and in the context of Omicron.

Overall, our study shows that an additional booster dose with mRNA vaccine had high effectiveness against severe COVID-19, independent of the COVID-19 vaccine scheme received in the past. Overall protection against COVID-19-related ICU admission and death showed a moderate decrease of about 9% and 6%, respectively, by the end of the follow-up. The introduction of the BA.4 and BA.5 Omicron sub-lineages during the second part of the study period (Supplementary Material Figure 3) and immunity waning due to a decline in the circulating levels of neutralizing antibodies as described for primary series and first booster doses, may explain the decrease in protection. We provide evidence to support mRNA-based second boosters following various background COVID-19 vaccine schemes, including widely used inactivated vaccines. These results suggest that an additional booster would help minimize severe COVID-19, including deaths, and decrease the impact on the health system. The observed duration of protection for a second booster dose suggests there may be longer-acting protection after repeated immunization with both homologous and heterologous schemes and that no additional boosters are needed at six months.

## Materials and Methods

### Exposures and outcomes

The Ministry of Health in Chile requires that all suspected COVID-19 cases are notified to health authorities through an online platform and undergo confirmatory laboratory testing. This is the source for the COVID-19 case count in this study. We assessed the effectiveness of mRNA-based second vaccine boosters for individuals with four different three-dose background regimes: (i) BNT162b2 primary series plus a homologous booster, (ii) CoronaVac primary series plus mRNA booster, (iii) CoronaVac primary series plus homologous booster, and (iv) CoronaVac primary series plus ChAdOx-1 booster. We estimated vaccine effectiveness weekly, from February 14, 2022, to August 15, 2022, against admission to an intensive care unit (ICU) and death (U07.1) associated with laboratory-confirmed SARS-CoV-2 infection.

We considered the onset of symptoms as a proxy for the time of infection. We used the time to the onset of symptoms from the beginning of the second booster campaign on February 14, 2022 as the endpoint of each outcome. Participants were classified into unvaccinated and fully immunized individuals (≥14 days after receipt of the additional mRNA booster dose). Individuals were excluded from the unvaccinated group when they received the first COVID-19 vaccine. We excluded the period between the first COVID-19 vaccine dose and 13 days after the additional mRNA booster dose from the at-risk person-time.

### Statistical analyses

Descriptive data was compared using Pearson’s χ^2^ tests. We estimated hazard ratios using an extension of the Cox hazards model to account for the time-varying vaccination status of participants.^21-23^ We estimated vaccine effectiveness using the hazard ratio between treated and non-treated individuals. Hazard ratios were adjusted for age, sex, region of residence, nationality, income, and comorbidities conditions (Supplementary Material Tables 1-5). We used a stratified version of the Cox hazards model^24^ with time-dependent covariates to compare the risk of the event of interest between immunized and non-immunized participants at each event time. Under the stratified Cox model, each combination of predictors has a specific hazard function that can evolve independently.

Let *T*_*i*_ be the time-to-event of interest, from February 14, 2022, for the *i*-th individual in the cohort, *i* = 1, …, *n*. Let *x*_*i*_, *i* = 1, …, *n*, be a *p-*dimensional vector of individual-specific characteristics, such as age and sex, and *z*_*i*_(*t*) be the time-dependent treatment indicator. The model assumes that the time-to-events are independent and with probability distribution given by

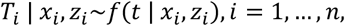

The time-to-event distribution is given by

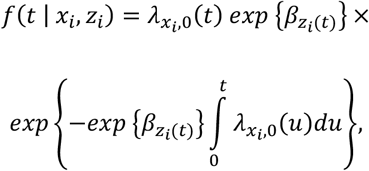

with *β*_*k*_ ∈ ℝ being the regression coefficient measuring the effectiveness of the *k*^th^ treatment, and λ_*x*,0_ is the predictor-specific baseline hazard function,

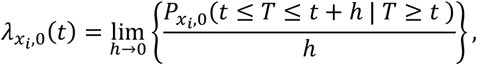

where 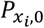 is the baseline probability distribution of the time-to-event in each strata.

We estimated the vaccine effectiveness as 100% · (1 − *exp* { *β*_*k*_}). We show the adjusted vaccine effectiveness results, including covariates as controls (age, gender, region, nationality, health insurance category, and comorbidities). We computed standard 95% Wald confidence intervals (95% CI) for the estimates. Inference was based on a partial likelihood approach.^25^ Each term in the partial likelihood of the effectiveness regression coefficient corresponds to the conditional probability of an individual expressing the outcome of interest from the risk set at a given calendar time.

We analyzed the data with the survival package^26^ of R, version 4.0.5.^27^

The Comité Ético Científico Clínica Alemana Universidad del Desarrollo, Santiago, Chile, gave ethical approval for this research. The study was considered exempt from informed consent.

## Data Availability

Owing to data privacy regulations in Chile, this study's individual-level data cannot be shared (Law 19.628). Aggregate data on vaccination, including demographics, and COVID-19 incidence, are publicly available at https://github.com/MinCiencia/Datos-Covid19/.

## Role of funders in the study design and execution

The funders of this study had no role in the study design, in the collection, analysis, and interpretation of data, in the writing of this manuscript or in the decision to submit the article for consideration for publication.

## Author contributions

AJ, CC, and RA conceived and designed the study. AJ, CC, MN, CG, and TB, and RA managed and analyzed the data. AJ, EU, RA wrote the first draft of the manuscript. AJ, CC, EU, MN, CG, MB, TB, and RA critically reviewed and edited the manuscript. VV, HE, and CC had access to vaccine safety data. All the authors are responsible for the study design, data collection, and data analysis. All authors have read and approved the final version of the manuscript. The authors vouch for the accuracy and completeness of the data and accept responsibility for publication. Drs. Jara, Cuadrado, and Araos contributed equally to the manuscript.

## Data availability

Owing to data privacy regulations in Chile, this study’s individual-level data cannot be shared (Law N19.628). Aggregate data on vaccination, including demographics, and COVID-19 incidence, are publicly available at https://github.com/MinCiencia/Datos-Covid19/.

## Supplementary Material

**Supplementary Material Table 1.**
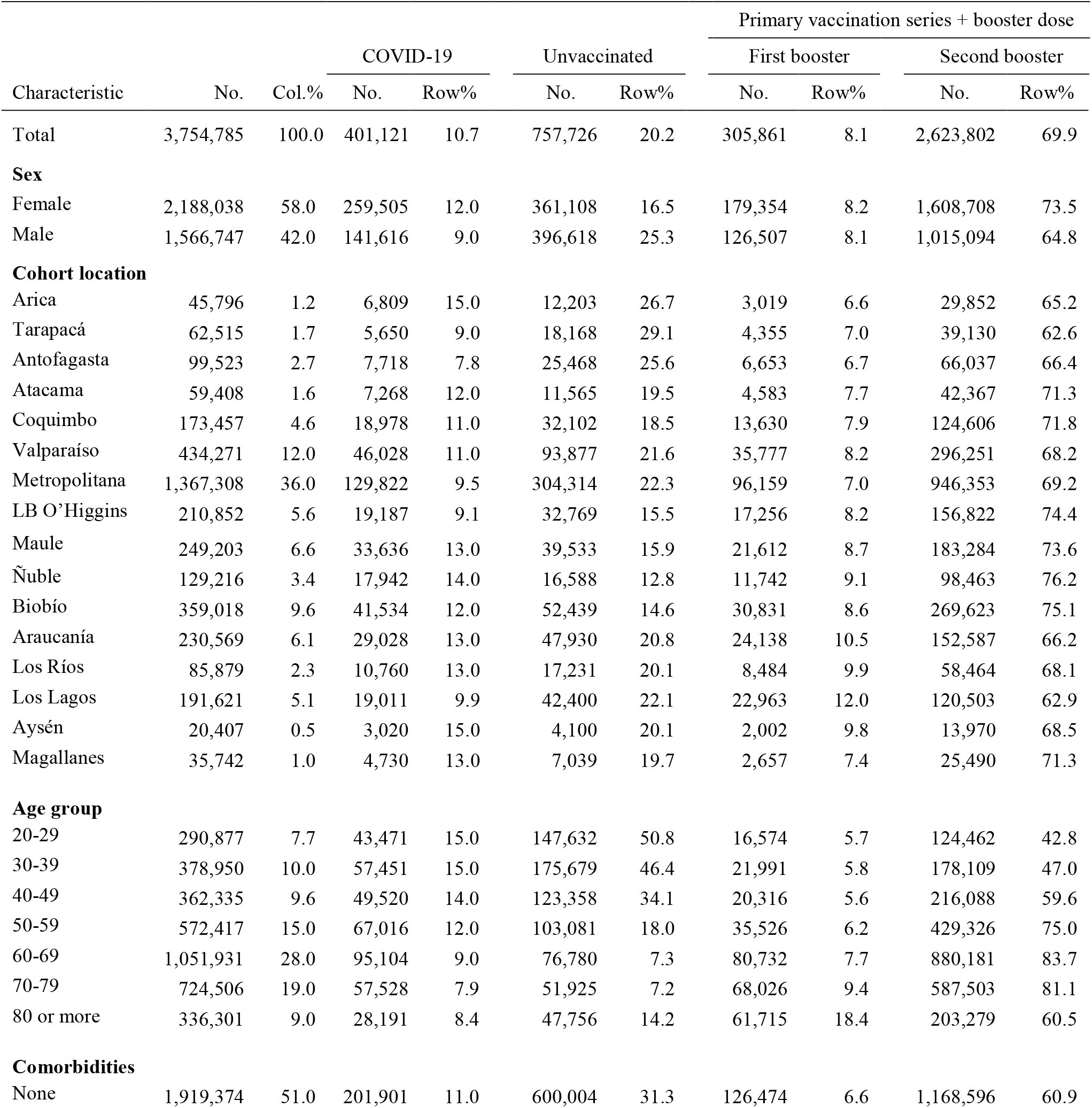
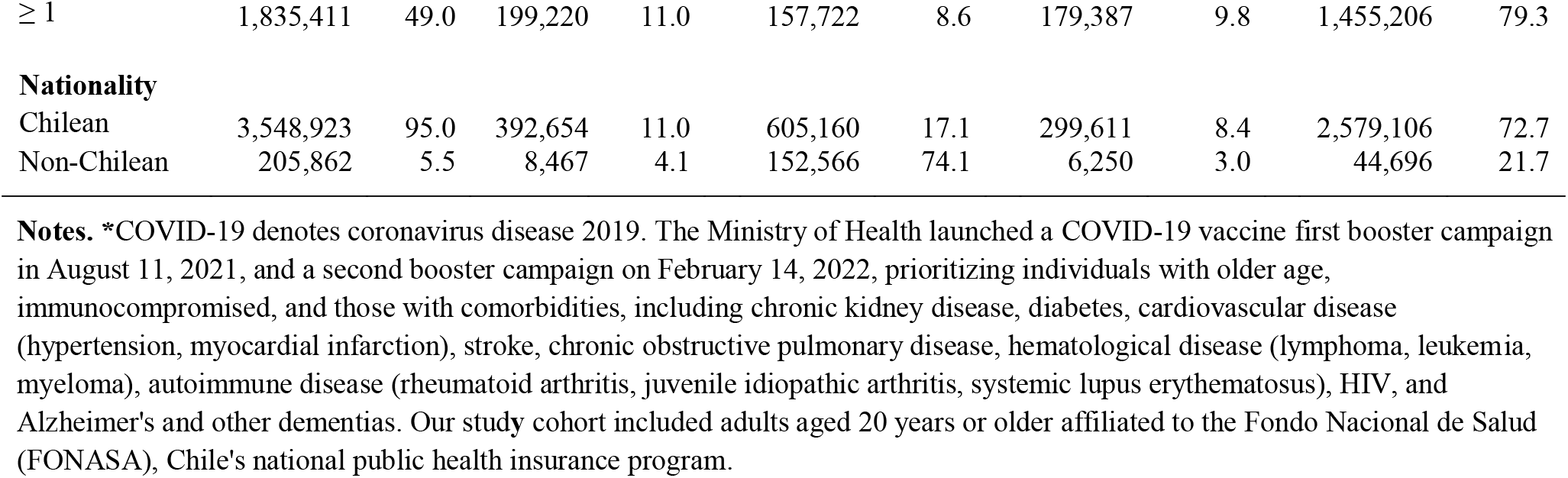
Characteristics of the study cohort of adults aged 20 years or older affiliated to FONASA, with laboratory-confirmed COVID-19, unvaccinated and vaccinated individuals who received three (booster) or four doses (additional booster) of COVID-19 vaccines, August 11, 2021, through August 18, 2022**\***

**Supplementary Material Table 2.**
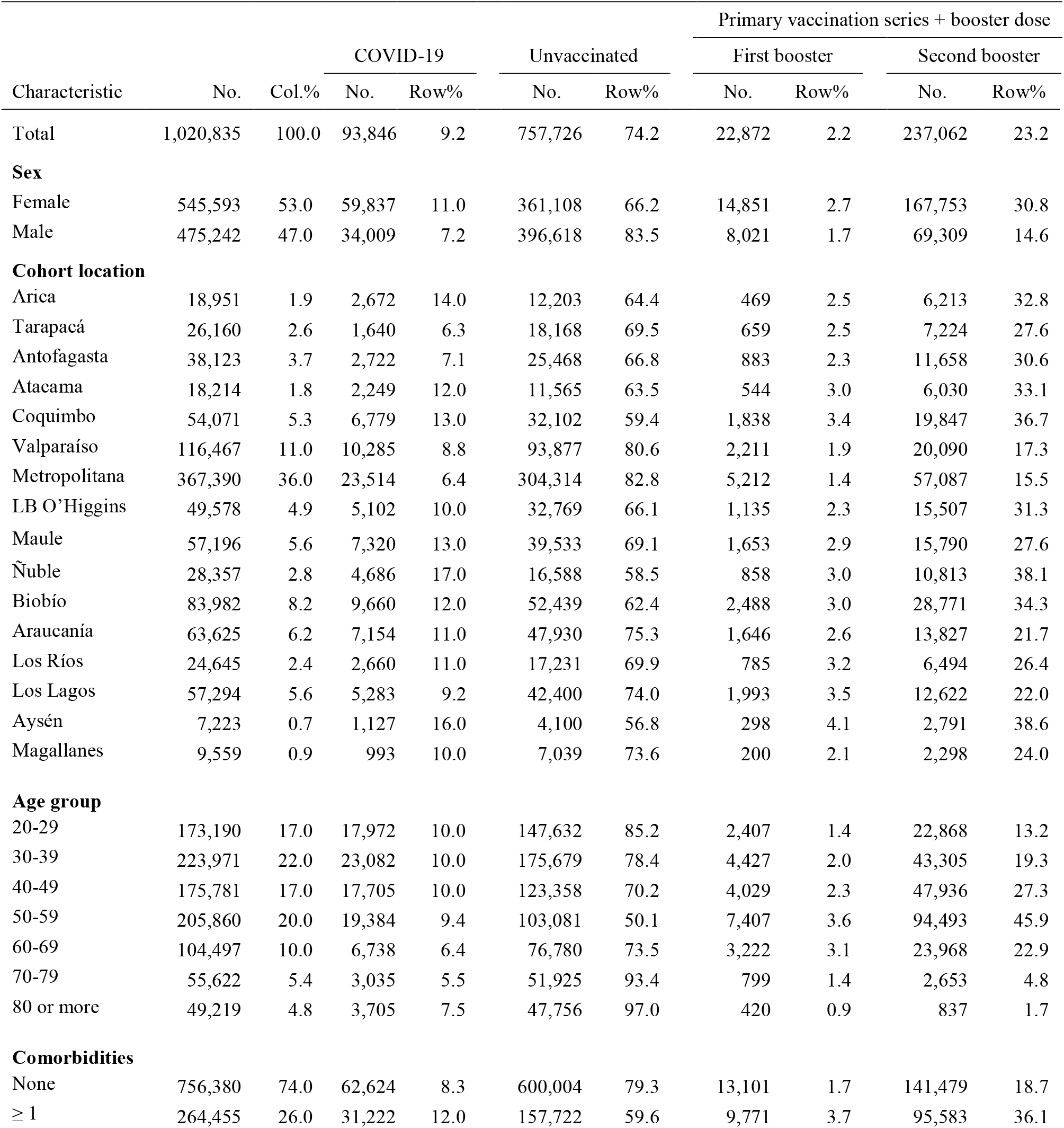
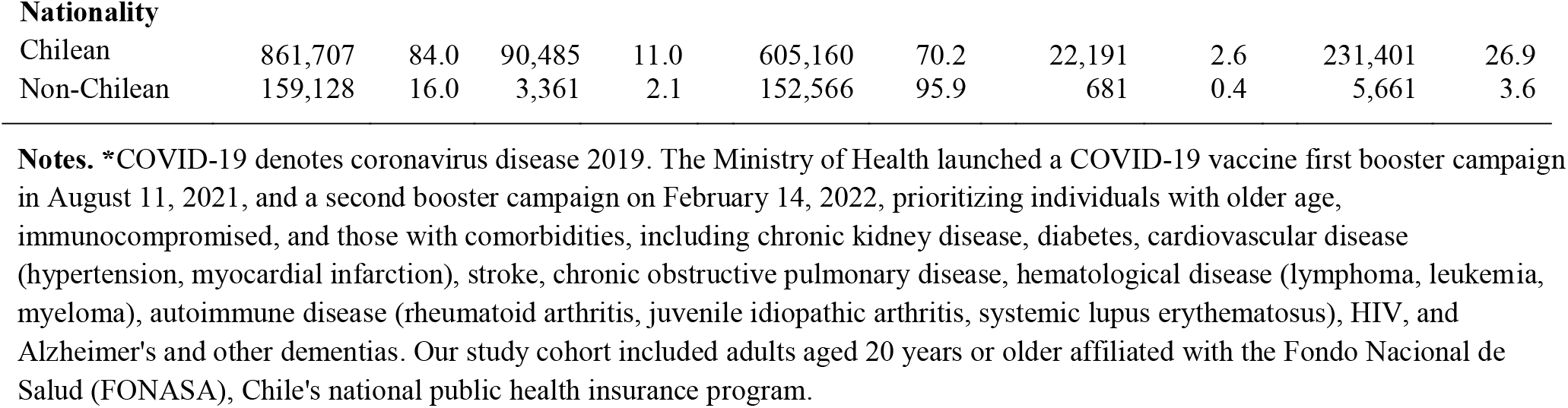
Characteristics of the study cohort of adults aged 20 years or older affiliated to FONASA, with laboratory-confirmed COVID-19, unvaccinated and vaccinated individuals who received a BNT162b2 primary series plus a homologous booster (3mRNA) and an additional mRNA booster (fourth dose), August 11, 2021, through August 18, 2022**\***

| Characteristic | No. | Col.% | COVID-19 |  | Unvaccinated |  | Primary vaccination series + booster dose |  |  |  |
| --- | --- | --- | --- | --- | --- | --- | --- | --- | --- | --- |
|  |  |  | No. | Row% | No. | Row% | First booster |  | Second booster |  |
|  |  |  |  |  |  |  | No. | Row% | No. | Row% |
| Total | 1,020,835 | 100.0 | 93,846 | 9.2 | 757,726 | 74.2 | 22,872 | 2.2 | 237,062 | 23.2 |
| **Sex** |  |  |  |  |  |  |  |  |  |  |
| Female | 545,593 | 53.0 | 59,837 | 11.0 | 361,108 | 66.2 | 14,851 | 2.7 | 167,753 | 30.8 |
| Male | 475,242 | 47.0 | 34,009 | 7.2 | 396,618 | 83.5 | 8,021 | 1.7 | 69,309 | 14.6 |
| **Cohort location** |  |  |  |  |  |  |  |  |  |  |
| Arica | 18,951 | 1.9 | 2,672 | 14.0 | 12,203 | 64.4 | 469 | 2.5 | 6,213 | 32.8 |
| Tarapacá | 26,160 | 2.6 | 1,640 | 6.3 | 18,168 | 69.5 | 659 | 2.5 | 7,224 | 27.6 |
| Antofagasta | 38,123 | 3.7 | 2,722 | 7.1 | 25,468 | 66.8 | 883 | 2.3 | 11,658 | 30.6 |
| Atacama | 18,214 | 1.8 | 2,249 | 12.0 | 11,565 | 63.5 | 544 | 3.0 | 6,030 | 33.1 |
| Coquimbo | 54,071 | 5.3 | 6,779 | 13.0 | 32,102 | 59.4 | 1,838 | 3.4 | 19,847 | 36.7 |
| Valparaíso | 116,467 | 11.0 | 10,285 | 8.8 | 93,877 | 80.6 | 2,211 | 1.9 | 20,090 | 17.3 |
| Metropolitana | 367,390 | 36.0 | 23,514 | 6.4 | 304,314 | 82.8 | 5,212 | 1.4 | 57,087 | 15.5 |
| LB O'Higgins | 49,578 | 4.9 | 5,102 | 10.0 | 32,769 | 66.1 | 1,135 | 2.3 | 15,507 | 31.3 |
| Maule | 57,196 | 5.6 | 7,320 | 13.0 | 39,533 | 69.1 | 1,653 | 2.9 | 15,790 | 27.6 |
| Ñuble | 28,357 | 2.8 | 4,686 | 17.0 | 16,588 | 58.5 | 858 | 3.0 | 10,813 | 38.1 |
| Biobío | 83,982 | 8.2 | 9,660 | 12.0 | 52,439 | 62.4 | 2,488 | 3.0 | 28,771 | 34.3 |
| Araucanía | 63,625 | 6.2 | 7,154 | 11.0 | 47,930 | 75.3 | 1,646 | 2.6 | 13,827 | 21.7 |
| Los Ríos | 24,645 | 2.4 | 2,660 | 11.0 | 17,231 | 69.9 | 785 | 3.2 | 6,494 | 26.4 |
| Los Lagos | 57,294 | 5.6 | 5,283 | 9.2 | 42,400 | 74.0 | 1,993 | 3.5 | 12,622 | 22.0 |
| Aysén | 7,223 | 0.7 | 1,127 | 16.0 | 4,100 | 56.8 | 298 | 4.1 | 2,791 | 38.6 |
| Magallanes | 9,559 | 0.9 | 993 | 10.0 | 7,039 | 73.6 | 200 | 2.1 | 2,298 | 24.0 |
| **Age group** |  |  |  |  |  |  |  |  |  |  |
| 20-29 | 173,190 | 17.0 | 17,972 | 10.0 | 147,632 | 85.2 | 2,407 | 1.4 | 22,868 | 13.2 |
| 30-39 | 223,971 | 22.0 | 23,082 | 10.0 | 175,679 | 78.4 | 4,427 | 2.0 | 43,305 | 19.3 |
| 40-49 | 175,781 | 17.0 | 17,705 | 10.0 | 123,358 | 70.2 | 4,029 | 2.3 | 47,936 | 27.3 |
| 50-59 | 205,860 | 20.0 | 19,384 | 9.4 | 103,081 | 50.1 | 7,407 | 3.6 | 94,493 | 45.9 |
| 60-69 | 104,497 | 10.0 | 6,738 | 6.4 | 76,780 | 73.5 | 3,222 | 3.1 | 23,968 | 22.9 |
| 70-79 | 55,622 | 5.4 | 3,035 | 5.5 | 51,925 | 93.4 | 799 | 1.4 | 2,653 | 4.8 |
| 80 or more | 49,219 | 4.8 | 3,705 | 7.5 | 47,756 | 97.0 | 420 | 0.9 | 837 | 1.7 |
| **Comorbidities** |  |  |  |  |  |  |  |  |  |  |
| None | 756,380 | 74.0 | 62,624 | 8.3 | 600,004 | 79.3 | 13,101 | 1.7 | 141,479 | 18.7 |
| ≥ 1 | 264,455 | 26.0 | 31,222 | 12.0 | 157,722 | 59.6 | 9,771 | 3.7 | 95,583 | 36.1 |
| **Nationality** |  |  |  |  |  |  |  |  |  |  |
| Chilean | 861,707 | 84.0 | 90,485 | 11.0 | 605,160 | 70.2 | 22,191 | 2.6 | 231,401 | 26.9 |
| Non-Chilean | 159,128 | 16.0 | 3,361 | 2.1 | 152,566 | 95.9 | 681 | 0.4 | 5,661 | 3.6 |

**Notes.** \*COVID-19 denotes coronavirus disease 2019. The Ministry of Health launched a COVID-19 vaccine first booster campaign in August 11, 2021, and a second booster campaign on February 14, 2022, prioritizing individuals with older age, immunocompromised, and those with comorbidities, including chronic kidney disease, diabetes, cardiovascular disease (hypertension, myocardial infarction), stroke, chronic obstructive pulmonary disease, hematological disease (lymphoma, leukemia, myeloma), autoimmune disease (rheumatoid arthritis, juvenile idiopathic arthritis, systemic lupus erythematosus), HIV, and Alzheimer's and other dementias. Our study cohort included adults aged 20 years or older affiliated with the Fondo Nacional de Salud (FONASA), Chile's national public health insurance program.

**Supplementary Material Table 3.**
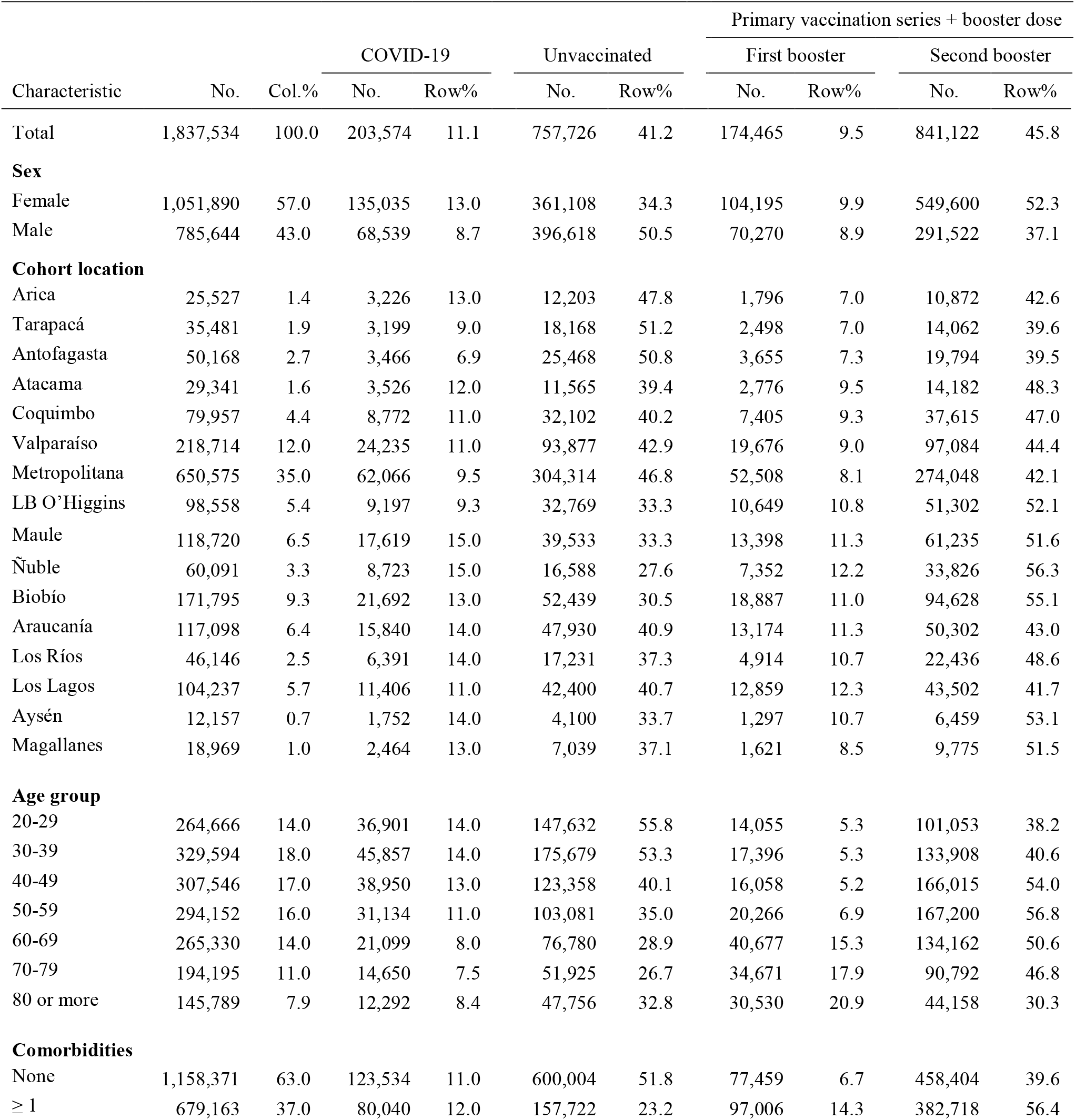
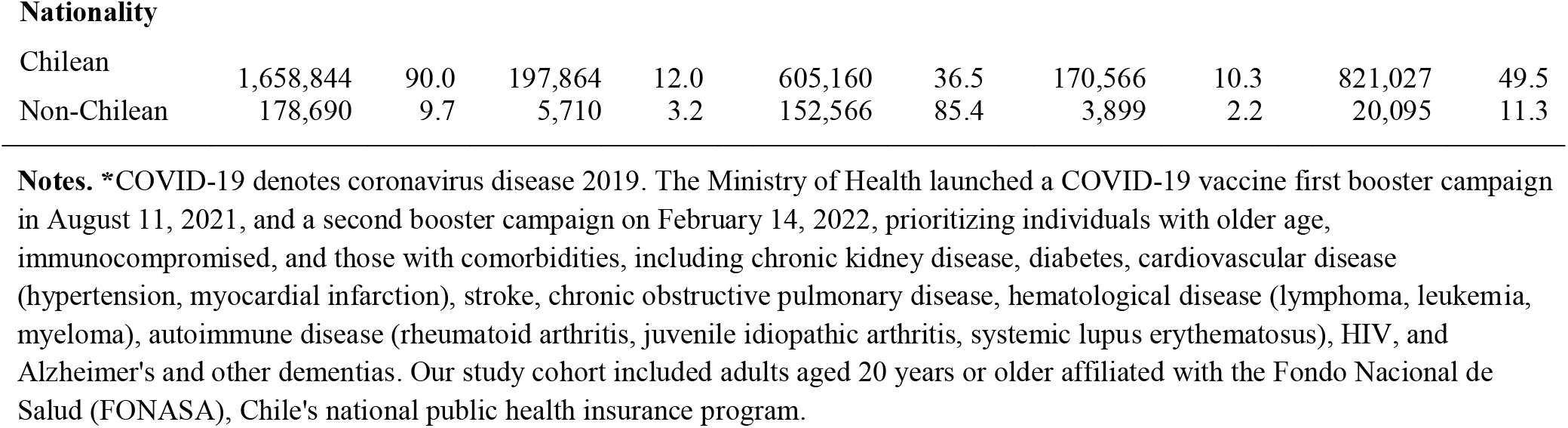
Characteristics of the study cohort of adults aged 20 years or older affiliated to FONASA, with laboratory-confirmed COVID-19, unvaccinated and vaccinated individuals who received an inactivated SARS-CoV-2 vaccine Sinovac primary series plus a heterologous mRNA booster (CC+mRNA) and an additional mRNA booster (fourth dose), August 11, 2021, through August 18, 2022**\***

| Characteristic | No. | Col.% | COVID-19 |  | Unvaccinated |  | Primary vaccination series + booster dose |  |  |  |
| --- | --- | --- | --- | --- | --- | --- | --- | --- | --- | --- |
|  |  |  | No. | Row% | No. | Row% | First booster |  | Second booster |  |
|  |  |  |  |  |  |  | No. | Row% | No. | Row% |
| Total | 1,837,534 | 100.0 | 203,574 | 11.1 | 757,726 | 41.2 | 174,465 | 9.5 | 841,122 | 45.8 |
| **Sex** |  |  |  |  |  |  |  |  |  |  |
| Female | 1,051,890 | 57.0 | 135,035 | 13.0 | 361,108 | 34.3 | 104,195 | 9.9 | 549,600 | 52.3 |
| Male | 785,644 | 43.0 | 68,539 | 8.7 | 396,618 | 50.5 | 70,270 | 8.9 | 291,522 | 37.1 |
| **Cohort location** |  |  |  |  |  |  |  |  |  |  |
| Arica | 25,527 | 1.4 | 3,226 | 13.0 | 12,203 | 47.8 | 1,796 | 7.0 | 10,872 | 42.6 |
| Tarapacá | 35,481 | 1.9 | 3,199 | 9.0 | 18,168 | 51.2 | 2,498 | 7.0 | 14,062 | 39.6 |
| Antofagasta | 50,168 | 2.7 | 3,466 | 6.9 | 25,468 | 50.8 | 3,655 | 7.3 | 19,794 | 39.5 |
| Atacama | 29,341 | 1.6 | 3,526 | 12.0 | 11,565 | 39.4 | 2,776 | 9.5 | 14,182 | 48.3 |
| Coquimbo | 79,957 | 4.4 | 8,772 | 11.0 | 32,102 | 40.2 | 7,405 | 9.3 | 37,615 | 47.0 |
| Valparaíso | 218,714 | 12.0 | 24,235 | 11.0 | 93,877 | 42.9 | 19,676 | 9.0 | 97,084 | 44.4 |
| Metropolitana | 650,575 | 35.0 | 62,066 | 9.5 | 304,314 | 46.8 | 52,508 | 8.1 | 274,048 | 42.1 |
| LB O'Higgins | 98,558 | 5.4 | 9,197 | 9.3 | 32,769 | 33.3 | 10,649 | 10.8 | 51,302 | 52.1 |
| Maule | 118,720 | 6.5 | 17,619 | 15.0 | 39,533 | 33.3 | 13,398 | 11.3 | 61,235 | 51.6 |
| Ñuble | 60,091 | 3.3 | 8,723 | 15.0 | 16,588 | 27.6 | 7,352 | 12.2 | 33,826 | 56.3 |
| Biobío | 171,795 | 9.3 | 21,692 | 13.0 | 52,439 | 30.5 | 18,887 | 11.0 | 94,628 | 55.1 |
| Araucanía | 117,098 | 6.4 | 15,840 | 14.0 | 47,930 | 40.9 | 13,174 | 11.3 | 50,302 | 43.0 |
| Los Ríos | 46,146 | 2.5 | 6,391 | 14.0 | 17,231 | 37.3 | 4,914 | 10.7 | 22,436 | 48.6 |
| Los Lagos | 104,237 | 5.7 | 11,406 | 11.0 | 42,400 | 40.7 | 12,859 | 12.3 | 43,502 | 41.7 |
| Aysén | 12,157 | 0.7 | 1,752 | 14.0 | 4,100 | 33.7 | 1,297 | 10.7 | 6,459 | 53.1 |
| Magallanes | 18,969 | 1.0 | 2,464 | 13.0 | 7,039 | 37.1 | 1,621 | 8.5 | 9,775 | 51.5 |
| **Age group** |  |  |  |  |  |  |  |  |  |  |
| 20-29 | 264,666 | 14.0 | 36,901 | 14.0 | 147,632 | 55.8 | 14,055 | 5.3 | 101,053 | 38.2 |
| 30-39 | 329,594 | 18.0 | 45,857 | 14.0 | 175,679 | 53.3 | 17,396 | 5.3 | 133,908 | 40.6 |
| 40-49 | 307,546 | 17.0 | 38,950 | 13.0 | 123,358 | 40.1 | 16,058 | 5.2 | 166,015 | 54.0 |
| 50-59 | 294,152 | 16.0 | 31,134 | 11.0 | 103,081 | 35.0 | 20,266 | 6.9 | 167,200 | 56.8 |
| 60-69 | 265,330 | 14.0 | 21,099 | 8.0 | 76,780 | 28.9 | 40,677 | 15.3 | 134,162 | 50.6 |
| 70-79 | 194,195 | 11.0 | 14,650 | 7.5 | 51,925 | 26.7 | 34,671 | 17.9 | 90,792 | 46.8 |
| 80 or more | 145,789 | 7.9 | 12,292 | 8.4 | 47,756 | 32.8 | 30,530 | 20.9 | 44,158 | 30.3 |
| **Comorbidities** |  |  |  |  |  |  |  |  |  |  |
| None | 1,158,371 | 63.0 | 123,534 | 11.0 | 600,004 | 51.8 | 77,459 | 6.7 | 458,404 | 39.6 |
| ≥ 1 | 679,163 | 37.0 | 80,040 | 12.0 | 157,722 | 23.2 | 97,006 | 14.3 | 382,718 | 56.4 |
| **Nationality** |  |  |  |  |  |  |  |  |  |  |
| Chilean | 1,658,844 | 90.0 | 197,864 | 12.0 | 605,160 | 36.5 | 170,566 | 10.3 | 821,027 | 49.5 |
| Non-Chilean | 178,690 | 9.7 | 5,710 | 3.2 | 152,566 | 85.4 | 3,899 | 2.2 | 20,095 | 11.3 |

**Notes.** \*COVID-19 denotes coronavirus disease 2019. The Ministry of Health launched a COVID-19 vaccine first booster campaign in August 11, 2021, and a second booster campaign on February 14, 2022, prioritizing individuals with older age, immunocompromised, and those with comorbidities, including chronic kidney disease, diabetes, cardiovascular disease (hypertension, myocardial infarction), stroke, chronic obstructive pulmonary disease, hematological disease (lymphoma, leukemia, myeloma), autoimmune disease (rheumatoid arthritis, juvenile idiopathic arthritis, systemic lupus erythematosus), HIV, and Alzheimer's and other dementias. Our study cohort included adults aged 20 years or older affiliated with the Fondo Nacional de Salud (FONASA), Chile's national public health insurance program.

**Supplementary Material Table 4.**
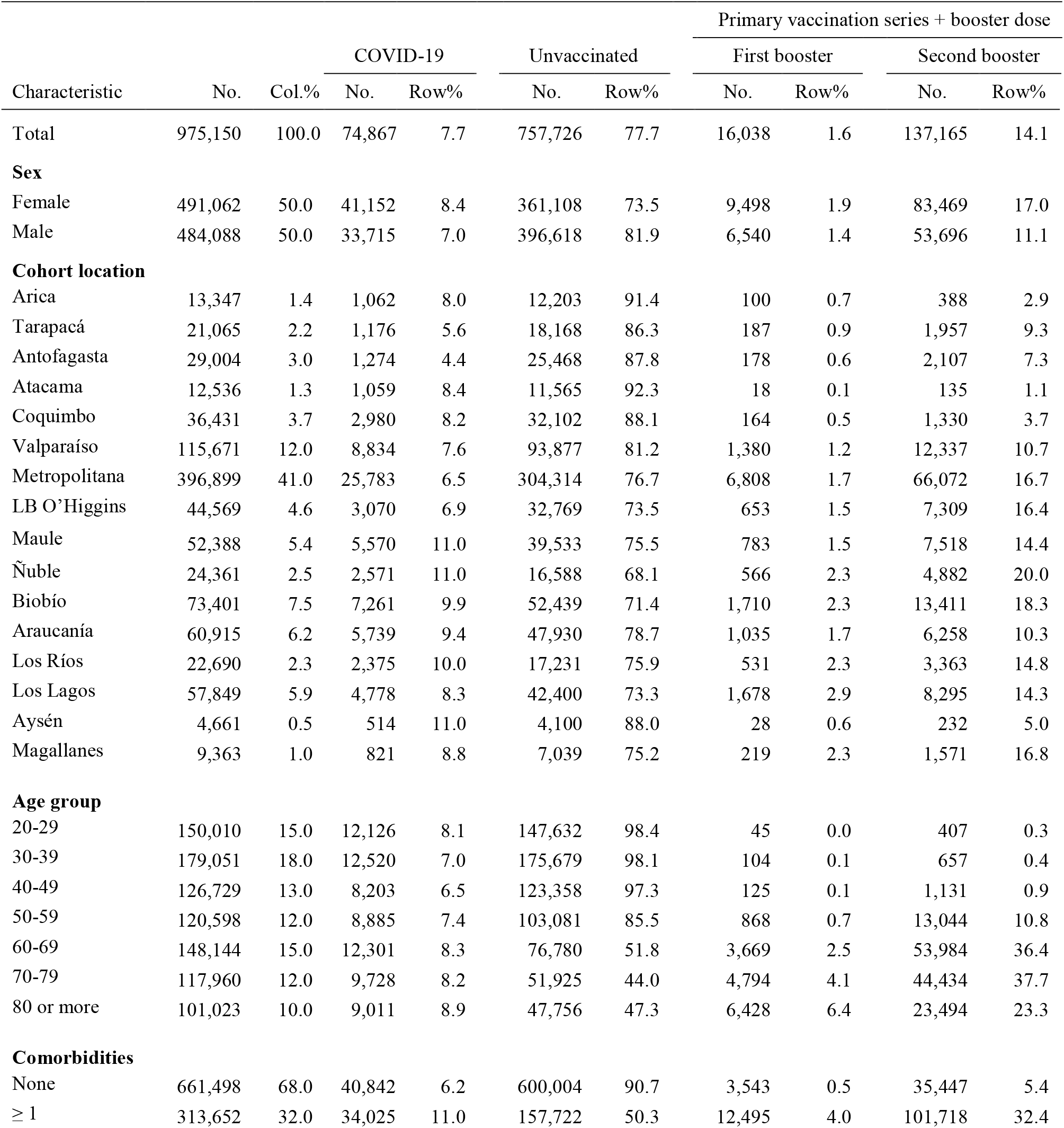
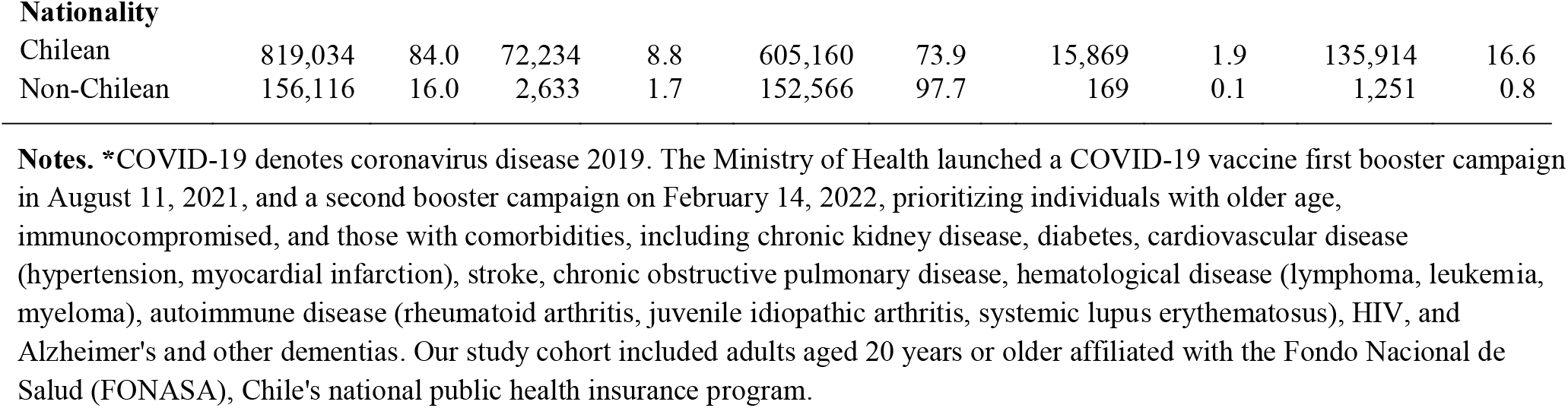
Characteristics of the study cohort of adults aged 20 years or older affiliated to FONASA, with laboratory-confirmed COVID-19, unvaccinated and vaccinated individuals who received an inactivated SARS-CoV-2 vaccine Sinovac primary series plus a homologous booster (CCC) and an additional mRNA booster (fourth dose), August 11, 2021, through August 18, 2022**\***

| Characteristic | No. | Col.% | COVID-19 |  | Unvaccinated |  | Primary vaccination series + booster dose |  |  |  |
| --- | --- | --- | --- | --- | --- | --- | --- | --- | --- | --- |
|  |  |  | No. | Row% | No. | Row% | First booster |  | Second booster |  |
|  |  |  |  |  |  |  | No. | Row% | No. | Row% |
| Total | 975,150 | 100.0 | 74,867 | 7.7 | 757,726 | 77.7 | 16,038 | 1.6 | 137,165 | 14.1 |
| **Sex** |  |  |  |  |  |  |  |  |  |  |
| Female | 491,062 | 50.0 | 41,152 | 8.4 | 361,108 | 73.5 | 9,498 | 1.9 | 83,469 | 17.0 |
| Male | 484,088 | 50.0 | 33,715 | 7.0 | 396,618 | 81.9 | 6,540 | 1.4 | 53,696 | 11.1 |
| **Cohort location** |  |  |  |  |  |  |  |  |  |  |
| Arica | 13,347 | 1.4 | 1,062 | 8.0 | 12,203 | 91.4 | 100 | 0.7 | 388 | 2.9 |
| Tarapacá | 21,065 | 2.2 | 1,176 | 5.6 | 18,168 | 86.3 | 187 | 0.9 | 1,957 | 9.3 |
| Antofagasta | 29,004 | 3.0 | 1,274 | 4.4 | 25,468 | 87.8 | 178 | 0.6 | 2,107 | 7.3 |
| Atacama | 12,536 | 1.3 | 1,059 | 8.4 | 11,565 | 92.3 | 18 | 0.1 | 135 | 1.1 |
| Coquimbo | 36,431 | 3.7 | 2,980 | 8.2 | 32,102 | 88.1 | 164 | 0.5 | 1,330 | 3.7 |
| Valparaíso | 115,671 | 12.0 | 8,834 | 7.6 | 93,877 | 81.2 | 1,380 | 1.2 | 12,337 | 10.7 |
| Metropolitana | 396,899 | 41.0 | 25,783 | 6.5 | 304,314 | 76.7 | 6,808 | 1.7 | 66,072 | 16.7 |
| LB O'Higgins | 44,569 | 4.6 | 3,070 | 6.9 | 32,769 | 73.5 | 653 | 1.5 | 7,309 | 16.4 |
| Maule | 52,388 | 5.4 | 5,570 | 11.0 | 39,533 | 75.5 | 783 | 1.5 | 7,518 | 14.4 |
| Ñuble | 24,361 | 2.5 | 2,571 | 11.0 | 16,588 | 68.1 | 566 | 2.3 | 4,882 | 20.0 |
| Biobío | 73,401 | 7.5 | 7,261 | 9.9 | 52,439 | 71.4 | 1,710 | 2.3 | 13,411 | 18.3 |
| Araucanía | 60,915 | 6.2 | 5,739 | 9.4 | 47,930 | 78.7 | 1,035 | 1.7 | 6,258 | 10.3 |
| Los Ríos | 22,690 | 2.3 | 2,375 | 10.0 | 17,231 | 75.9 | 531 | 2.3 | 3,363 | 14.8 |
| Los Lagos | 57,849 | 5.9 | 4,778 | 8.3 | 42,400 | 73.3 | 1,678 | 2.9 | 8,295 | 14.3 |
| Aysén | 4,661 | 0.5 | 514 | 11.0 | 4,100 | 88.0 | 28 | 0.6 | 232 | 5.0 |
| Magallanes | 9,363 | 1.0 | 821 | 8.8 | 7,039 | 75.2 | 219 | 2.3 | 1,571 | 16.8 |
| **Age group** |  |  |  |  |  |  |  |  |  |  |
| 20-29 | 150,010 | 15.0 | 12,126 | 8.1 | 147,632 | 98.4 | 45 | 0.0 | 407 | 0.3 |
| 30-39 | 179,051 | 18.0 | 12,520 | 7.0 | 175,679 | 98.1 | 104 | 0.1 | 657 | 0.4 |
| 40-49 | 126,729 | 13.0 | 8,203 | 6.5 | 123,358 | 97.3 | 125 | 0.1 | 1,131 | 0.9 |
| 50-59 | 120,598 | 12.0 | 8,885 | 7.4 | 103,081 | 85.5 | 868 | 0.7 | 13,044 | 10.8 |
| 60-69 | 148,144 | 15.0 | 12,301 | 8.3 | 76,780 | 51.8 | 3,669 | 2.5 | 53,984 | 36.4 |
| 70-79 | 117,960 | 12.0 | 9,728 | 8.2 | 51,925 | 44.0 | 4,794 | 4.1 | 44,434 | 37.7 |
| 80 or more | 101,023 | 10.0 | 9,011 | 8.9 | 47,756 | 47.3 | 6,428 | 6.4 | 23,494 | 23.3 |
| **Comorbidities** |  |  |  |  |  |  |  |  |  |  |
| None | 661,498 | 68.0 | 40,842 | 6.2 | 600,004 | 90.7 | 3,543 | 0.5 | 35,447 | 5.4 |
| ≥ 1 | 313,652 | 32.0 | 34,025 | 11.0 | 157,722 | 50.3 | 12,495 | 4.0 | 101,718 | 32.4 |
| **Nationality** |  |  |  |  |  |  |  |  |  |  |
| Chilean | 819,034 | 84.0 | 72,234 | 8.8 | 605,160 | 73.9 | 15,869 | 1.9 | 135,914 | 16.6 |
| Non-Chilean | 156,116 | 16.0 | 2,633 | 1.7 | 152,566 | 97.7 | 169 | 0.1 | 1,251 | 0.8 |

**Notes.** \*COVID-19 denotes coronavirus disease 2019. The Ministry of Health launched a COVID-19 vaccine first booster campaign in August 11, 2021, and a second booster campaign on February 14, 2022, prioritizing individuals with older age, immunocompromised, and those with comorbidities, including chronic kidney disease, diabetes, cardiovascular disease (hypertension, myocardial infarction), stroke, chronic obstructive pulmonary disease, hematological disease (lymphoma, leukemia, myeloma), autoimmune disease (rheumatoid arthritis, juvenile idiopathic arthritis, systemic lupus erythematosus), HIV, and Alzheimer's and other dementias. Our study cohort included adults aged 20 years or older affiliated with the Fondo Nacional de Salud (FONASA), Chile's national public health insurance program.

**Supplementary Material Table 5.**
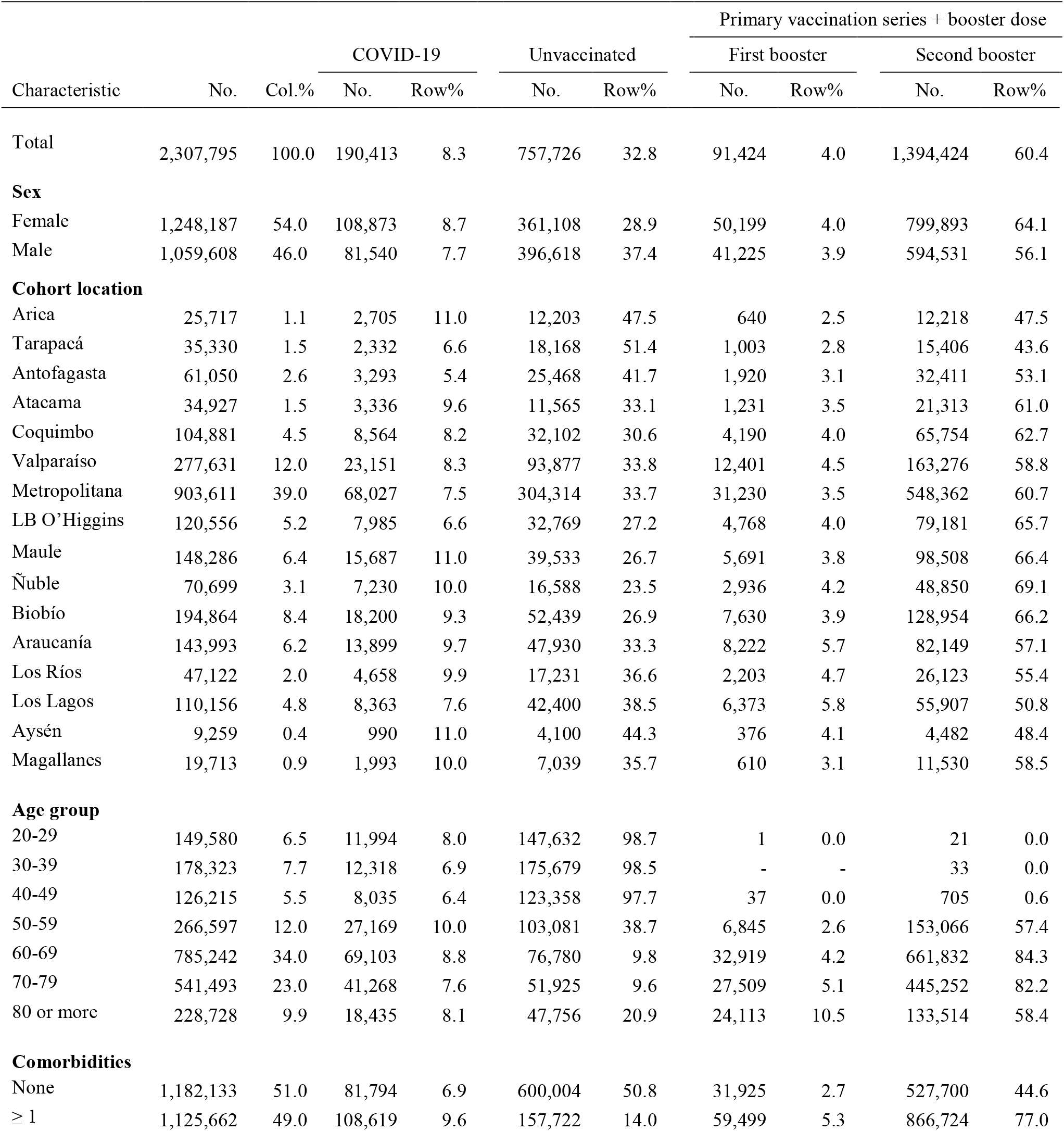
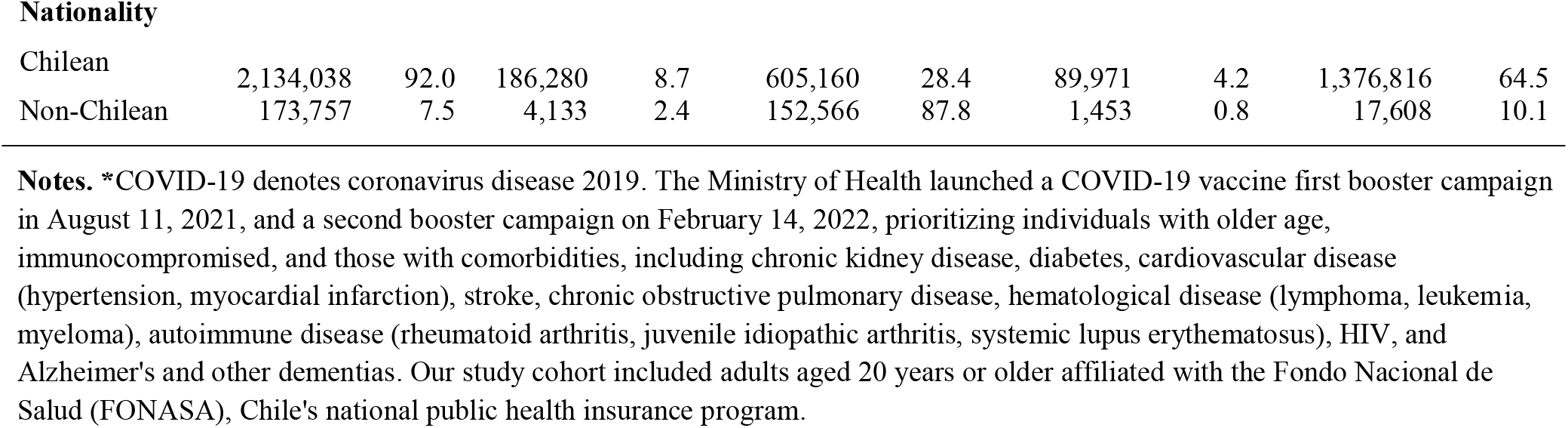
Characteristics of the study cohort of adults aged 20 years or older affiliated to FONASA, with laboratory-confirmed COVID-19, unvaccinated and vaccinated individuals who received an inactivated SARS-CoV-2 vaccine Sinovac primary series plus a heterologous viral-vectored ChAdOx1 booster (CCA) and an additional mRNA booster (fourth dose), August 11, 2021, through August 18, 2022**\***

| Characteristic | No. | Col.% | COVID-19 |  | Unvaccinated |  | Primary vaccination series + booster dose |  |  |  |
| --- | --- | --- | --- | --- | --- | --- | --- | --- | --- | --- |
|  |  |  | No. | Row% | No. | Row% | First booster |  | Second booster |  |
|  |  |  |  |  |  |  | No. | Row% | No. | Row% |
| Total | 2,307,795 | 100.0 | 190,413 | 8.3 | 757,726 | 32.8 | 91,424 | 4.0 | 1,394,424 | 60.4 |
| **Sex** |  |  |  |  |  |  |  |  |  |  |
| Female | 1,248,187 | 54.0 | 108,873 | 8.7 | 361,108 | 28.9 | 50,199 | 4.0 | 799,893 | 64.1 |
| Male | 1,059,608 | 46.0 | 81,540 | 7.7 | 396,618 | 37.4 | 41,225 | 3.9 | 594,531 | 56.1 |
| **Cohort location** |  |  |  |  |  |  |  |  |  |  |
| Arica | 25,717 | 1.1 | 2,705 | 11.0 | 12,203 | 47.5 | 640 | 2.5 | 12,218 | 47.5 |
| Tarapacá | 35,330 | 1.5 | 2,332 | 6.6 | 18,168 | 51.4 | 1,003 | 2.8 | 15,406 | 43.6 |
| Antofagasta | 61,050 | 2.6 | 3,293 | 5.4 | 25,468 | 41.7 | 1,920 | 3.1 | 32,411 | 53.1 |
| Atacama | 34,927 | 1.5 | 3,336 | 9.6 | 11,565 | 33.1 | 1,231 | 3.5 | 21,313 | 61.0 |
| Coquimbo | 104,881 | 4.5 | 8,564 | 8.2 | 32,102 | 30.6 | 4,190 | 4.0 | 65,754 | 62.7 |
| Valparaíso | 277,631 | 12.0 | 23,151 | 8.3 | 93,877 | 33.8 | 12,401 | 4.5 | 163,276 | 58.8 |
| Metropolitana | 903,611 | 39.0 | 68,027 | 7.5 | 304,314 | 33.7 | 31,230 | 3.5 | 548,362 | 60.7 |
| LB O'Higgins | 120,556 | 5.2 | 7,985 | 6.6 | 32,769 | 27.2 | 4,768 | 4.0 | 79,181 | 65.7 |
| Maule | 148,286 | 6.4 | 15,687 | 11.0 | 39,533 | 26.7 | 5,691 | 3.8 | 98,508 | 66.4 |
| Ñuble | 70,699 | 3.1 | 7,230 | 10.0 | 16,588 | 23.5 | 2,936 | 4.2 | 48,850 | 69.1 |
| Biobío | 194,864 | 8.4 | 18,200 | 9.3 | 52,439 | 26.9 | 7,630 | 3.9 | 128,954 | 66.2 |
| Araucanía | 143,993 | 6.2 | 13,899 | 9.7 | 47,930 | 33.3 | 8,222 | 5.7 | 82,149 | 57.1 |
| Los Ríos | 47,122 | 2.0 | 4,658 | 9.9 | 17,231 | 36.6 | 2,203 | 4.7 | 26,123 | 55.4 |
| Los Lagos | 110,156 | 4.8 | 8,363 | 7.6 | 42,400 | 38.5 | 6,373 | 5.8 | 55,907 | 50.8 |
| Aysén | 9,259 | 0.4 | 990 | 11.0 | 4,100 | 44.3 | 376 | 4.1 | 4,482 | 48.4 |
| Magallanes | 19,713 | 0.9 | 1,993 | 10.0 | 7,039 | 35.7 | 610 | 3.1 | 11,530 | 58.5 |
| **Age group** |  |  |  |  |  |  |  |  |  |  |
| 20-29 | 149,580 | 6.5 | 11,994 | 8.0 | 147,632 | 98.7 | 1 | 0.0 | 21 | 0.0 |
| 30-39 | 178,323 | 7.7 | 12,318 | 6.9 | 175,679 | 98.5 | - | - | 33 | 0.0 |
| 40-49 | 126,215 | 5.5 | 8,035 | 6.4 | 123,358 | 97.7 | 37 | 0.0 | 705 | 0.6 |
| 50-59 | 266,597 | 12.0 | 27,169 | 10.0 | 103,081 | 38.7 | 6,845 | 2.6 | 153,066 | 57.4 |
| 60-69 | 785,242 | 34.0 | 69,103 | 8.8 | 76,780 | 9.8 | 32,919 | 4.2 | 661,832 | 84.3 |
| 70-79 | 541,493 | 23.0 | 41,268 | 7.6 | 51,925 | 9.6 | 27,509 | 5.1 | 445,252 | 82.2 |
| 80 or more | 228,728 | 9.9 | 18,435 | 8.1 | 47,756 | 20.9 | 24,113 | 10.5 | 133,514 | 58.4 |
| **Comorbidities** |  |  |  |  |  |  |  |  |  |  |
| None | 1,182,133 | 51.0 | 81,794 | 6.9 | 600,004 | 50.8 | 31,925 | 2.7 | 527,700 | 44.6 |
| ≥ 1 | 1,125,662 | 49.0 | 108,619 | 9.6 | 157,722 | 14.0 | 59,499 | 5.3 | 866,724 | 77.0 |
| **Nationality** |  |  |  |  |  |  |  |  |  |  |
| Chilean | 2,134,038 | 92.0 | 186,280 | 8.7 | 605,160 | 28.4 | 89,971 | 4.2 | 1,376,816 | 64.5 |
| Non-Chilean | 173,757 | 7.5 | 4,133 | 2.4 | 152,566 | 87.8 | 1,453 | 0.8 | 17,608 | 10.1 |

**Notes.** \*COVID-19 denotes coronavirus disease 2019. The Ministry of Health launched a COVID-19 vaccine first booster campaign in August 11, 2021, and a second booster campaign on February 14, 2022, prioritizing individuals with older age, immunocompromised, and those with comorbidities, including chronic kidney disease, diabetes, cardiovascular disease (hypertension, myocardial infarction), stroke, chronic obstructive pulmonary disease, hematological disease (lymphoma, leukemia, myeloma), autoimmune disease (rheumatoid arthritis, juvenile idiopathic arthritis, systemic lupus erythematosus), HIV, and Alzheimer's and other dementias. Our study cohort included adults aged 20 years or older affiliated with the Fondo Nacional de Salud (FONASA), Chile's national public health insurance program.

**Supplementary Material Table 6.** Weekly effectiveness against ICU admissions of mRNA-based second vaccine boosters for individuals with four different three-dose background regimes: (i) BNT162b2 primary series plus a homologous booster, a CoronaVac primary series plus (ii) mRNA booster, (iii) homologous booster, or (iv) ChAdOx-1 booster, compared to unvaccinated individuals among adults aged 20 years and older, March 7, 2022, through August 15, 2022*

| Report date | Overall |  | 3mRNA+mRNA |  | CC+mRNA+mRNA |  | CCC+mRNA |  | CCA+mRNA |  |
| --- | --- | --- | --- | --- | --- | --- | --- | --- | --- | --- |
|  | VE (%) | 95% CI | VE (%) | 95% CI | VE (%) | 95% CI | VE (%) | 95% CI | VE (%) | 95% CI |
| March 7 | 96.8 | (86.8-99.2) | 100 |  | 100.0 |  | 88.0 | (13.6-98.3) | 95.6 | (81.9-99) |
| March 14 | 94.1 | (88.3-97.0) | 100 |  | 93.0 | (49.6-99.0) | 93.6 | (53.6-99.1) | 94.7 | (87.7-97.7) |
| March 21 | 96.3 | (92.9-98.1) | 100 |  | 92.0 | (66.6-98.0) | 95.0 | (64.0-99.3) | 96.9 | (92.9-98.7) |
| March 28 | 95.9 | (92.9-97.6) | 100 |  | 90.0 | (68.6-96.9) | 92.2 | (68.1-98.1) | 96.3 | (92.7-98.1) |
| April 4 | 95.7 | (93.1-97.3) | 100 |  | 87.0 | (67.2-94.6) | 92.9 | (71.2-98.3) | 96.4 | (93.2-98.1) |
| April 11 | 95.5 | (93.1-97.1) | 100 |  | 83.0 | (63.6-92.2) | 93.7 | (74.5-98.5) | 96.1 | (93.0-97.8) |
| April 18 | 96.0 | (93.8-97.4) | 100 |  | 85.0 | (67.4-93.0) | 91.2 | (72.2-97.2) | 96.5 | (93.7-98.0) |
| April 25 | 96.0 | (94.0-97.4) | 100 |  | 86.0 | (69.4-93.4) | 88.7 | (69.3-95.8) | 96.6 | (94.0-98.1) |
| May 2 | 95.7 | (93.6-97.1) | 100 |  | 86.0 | (70.4-93.6) | 89.6 | (71.8-96.2) | 96.3 | (93.6-97.8) |
| May 9 | 95.5 | (93.5-96.9) | 100 |  | 88.0 | (73.3-94.2) | 85.2 | (66.3-93.5) | 95.7 | (92.9-97.3) |
| May 16 | 94.9 | (92.8-96.4) | 100 |  | 88.0 | (74.6-94.5) | 85.9 | (67.8-93.8) | 95.0 | (92.2-96.8) |
| May 23 | 94.5 | (92.4-96.0) | 100 |  | 84.0 | (69.9-91.8) | 87.2 | (70.9-94.4) | 94.2 | (91.2-96.2) |
| May 30 | 94.2 | (92.1-95.7) | 85.5 | (0.0-98.0) | 85.0 | (71.9-91.9) | 88.7 | (74.2-95.0) | 93.5 | (90.5-95.5) |
| June 6 | 93.1 | (91.0-94.7) | 70.8 | (5.6-91.0) | 84.0 | (72.0-90.8) | 82.5 | (67.5-90.6) | 92.4 | (89.4-94.6) |
| June 13 | 92.3 | (90.2-93.9) | 74.2 | (28.4-90.7) | 80.0 | (68.8-87.5) | 77.7 | (62.5-86.8) | 92.1 | (89.2-94.2) |
| June 20 | 91.8 | (89.8-93.4) | 75.9 | (39.8-90.3) | 80.0 | (69.4-86.8) | 75.9 | (61.0-85.1) | 91.4 | (88.6-93.6) |
| June 27 | 90.8 | (88.7-92.5) | 75.8 | (44.0-89.5) | 78.0 | (68.2-85.3) | 78.6 | (65.4-86.7) | 89.8 | (86.7-92.2) |
| July 4 | 90.7 | (88.8-92.3) | 77.2 | (50.4-89.5) | 78.0 | (68.2-84.4) | 78.0 | (65.2-86.0) | 89.7 | (86.7-92.0) |
| July 11 | 89.7 | (87.7-91.4) | 79.9 | (56.4-90.7) | 78.0 | (68.6-84.1) | 74.8 | (61.5-83.5) | 87.9 | (84.8-90.4) |
| July 18 | 89.3 | (87.3-91.0) | 75.8 | (51.8-87.8) | 78.0 | (69.0-84.1) | 72.9 | (59.4-81.9) | 87.1 | (83.9-89.7) |
| July 25 | 88.5 | (86.4-90.2) | 72.4 | (48.3-85.3) | 77.0 | (68.3-83.4) | 74.3 | (61.9-82.7) | 86.0 | (82.6-88.7) |
| August 01 | 88.5 | (86.4-90.2) | 74.2 | (53.0-85.9) | 77.0 | (69.1-83.3) | 74.4 | (62.5-82.5) | 86.3 | (83.1-88.9) |
| August 08 | 88.3 | (86.3-90.0) | 75.0 | (55.4-86.0) | 77.0 | (68.7-82.6) | 75.0 | (63.4-82.9) | 86.7 | (83.6-89.2) |
| August 15 | 88.2 | (86.2-89.9) | 74.2 | (54.8-85.2) | 77.0 | (69.7-83.0) | 74.2 | (54.8-85.2) | 86.3 | (83.2-88.8) |

\*COVID-19 denotes coronavirus disease 2019. The Ministry of Health launched a COVID-19 vaccine first booster campaign in August 11, 2021, and a second booster campaign on February 14, 2022. The table shows the estimated vaccine effectiveness of mRNA-based second vaccine boosters for individuals with four different three-dose background regimes: i) BNT162b2 primary series plus a homologous booster (3mRNA), a CoronaVac primary series plus (ii) mRNA booster (CC+mRNA), iii) homologous booster (CCC), or (iv) ChAdOx-1 booster (CCA), compared to unvaccinated individuals. Estimates were adjusted for time-varying vaccination exposure and clinical, demographic, and socioeconomic confounders at baseline (Supplementary Material Tables 1-5).

**Supplementary Material Table 7.** Weekly effectiveness against confirmed death of mRNA-based second vaccine boosters for individuals with four different three-dose background regimes: (i) BNT162b2 primary series plus a homologous booster, a CoronaVac primary series plus mRNA booster, (iii) homologous booster, or (iv) ChAdOx-1 booster, compared to unvaccinated individuals among adults aged 20 years and older, March 7, 2022, through August 15, 2022*

| Report date | Overall |  | 3mRNA+mRNA |  | CC+mRNA+mRNA |  | CCC+mRNA |  | CCA+mRNA |  |
| --- | --- | --- | --- | --- | --- | --- | --- | --- | --- | --- |
|  | VE (%) | 95% CI | VE (%) | 95% CI | VE (%) | 95% CI | VE (%) | 95% CI | VE (%) | 95% CI |
| March 7 | 96.5 | (93.0-98.3) | 100.0 |  | 95.0 | (64.3-99.3) | 79.6 | (17.2-95.0) | 96.5 | (91.3-98.6) |
| March 14 | 96.6 | (94.6-97.9) | 100.0 |  | 98.0 | (86.3-99.7) | 89.4 | (66.7-96.6) | 96.2 | (93.4-97.8) |
| March 21 | 96.9 | (95.4-97.9) | 100.0 |  | 98.0 | (90.1-99.4) | 93.6 | (80.1-98.0) | 96.5 | (94.4-97.8) |
| March 28 | 96.7 | (95.3-97.7) | 100.0 |  | 97.0 | (91.1-99.1) | 93.8 | (83.2-97.7) | 96.5 | (94.7-97.6) |
| April 4 | 96.5 | (95.3-97.4) | 100.0 |  | 96.0 | (90.6-98.4) | 91.3 | (81.6-95.9) | 96.4 | (94.8-97.5) |
| April 11 | 96.6 | (95.5-97.5) | 100.0 |  | 97.0 | (91.6-98.6) | 92.4 | (83.8-96.4) | 96.5 | (95.0-97.5) |
| April 18 | 96.6 | (95.5-97.5) | 100.0 |  | 96.0 | (91.3-98.3) | 92.9 | (85.0-96.7) | 96.4 | (95.0-97.4) |
| April 25 | 96.4 | (95.3-97.3) | 100.0 |  | 95.0 | (90.2-97.6) | 90.6 | (82.3-95.0) | 96.4 | (95.0-97.4) |
| May 2 | 96.2 | (95.1-97.1) | 100.0 |  | 95.0 | (90.5-97.7) | 90.0 | (81.8-94.5) | 96.2 | (94.8-97.2) |
| May 9 | 95.8 | (94.7-96.7) | 100.0 |  | 96.0 | (91.2-97.8) | 89.1 | (80.9-93.7) | 95.7 | (94.3-96.8) |
| May 16 | 95.5 | (94.4-96.4) | 100.0 |  | 95.0 | (90.9-97.6) | 88.8 | (80.9-93.5) | 95.2 | (93.7-96.4) |
| May 23 | 95.2 | (94.1-96.1) | 100.0 |  | 94.0 | (89.2-96.4) | 88.2 | (80.4-92.8) | 95.0 | (93.5-96.1) |
| May 30 | 95.0 | (93.9-95.9) | 100.0 |  | 93.0 | (87.9-95.5) | 87.7 | (80.2-92.4) | 94.8 | (93.3-95.9) |
| June 6 | 94.1 | (92.9-95.0) | 100.0 |  | 90.0 | (84.8-93.1) | 84.8 | (77.1-89.9) | 94.0 | (92.5-95.2) |
| June 13 | 93.4 | (92.3-94.4) | 100.0 |  | 87.0 | (82.0-90.8) | 85.1 | (78.0-90.0) | 93.2 | (91.7-94.5) |
| June 20 | 92.9 | (91.8-93.9) | 100.0 |  | 86.0 | (81.0-89.5) | 82.8 | (75.8-87.8) | 92.8 | (91.3-94.1) |
| June 27 | 92.5 | (91.4-93.4) | 100.0 | (73.3-96.3) | 86.0 | (81.8-89.6) | 81.0 | (74.0-86.0) | 92.2 | (90.7-93.5) |
| July 4 | 92.1 | (91.0-93.0) | 90.1 | (76.0-95.9) | 84.0 | (79.8-87.6) | 81.4 | (75.0-86.1) | 91.9 | (90.5-93.2) |
| July 11 | 91.5 | (90.4-92.4) | 90.1 | (75.7-95.2) | 83.0 | (79.3-86.8) | 80.1 | (74.0-84.8) | 91.5 | (90.0-92.7) |
| July 18 | 91.0 | (89.9-92.0) | 89.2 | (75.7-94.6) | 83.0 | (78.7-86.1) | 78.2 | (72.0-83.1) | 91.0 | (89.6-92.3) |
| July 25 | 90.6 | (89.6-91.6) | 88.6 | (74.2-93.2) | 82.0 | (77.9-85.3) | 78.5 | (72.5-83.2) | 90.6 | (89.1-91.8) |
| August 01 | 90.5 | (89.5-91.5) | 86.7 | (75.8-93.6) | 81.0 | (77.0-84.4) | 79.3 | (73.6-83.7) | 90.7 | (89.3-91.9) |
| August 08 | 90.6 | (89.5-91.5) | 87.6 | (76.1-93.7) | 81.0 | (77.3-84.4) | 79.7 | (74.1-84.0) | 90.9 | (89.5-92.1) |
| August 15 | 90.5 | (89.4-91.4) | 87.7 | (69.7-83.0) | 81.0 | (76.8-84.0) | 79.3 | (73.8-83.7) | 90.8 | (89.4-92.0) |

\*COVID-19 denotes coronavirus disease 2019. The Ministry of Health launched a COVID-19 vaccine first booster campaign in August 11, 2021, and a second booster campaign on February 14, 2022. The table shows the estimated vaccine effectiveness of mRNA-based second vaccine boosters for individuals with four different three-dose background regimes: i) BNT162b2 primary series plus a homologous booster (3mRNA), a CoronaVac primary series plus (ii) mRNA booster (CC+mRNA), iii) homologous booster (CCC), or (iv) ChAdOx-1 booster (CCA), compared to unvaccinated individuals. Estimates were adjusted for time-varying vaccination exposure and clinical, demographic, and socioeconomic confounders at baseline (Supplementary Material Tables 1-5).

**Supplementary Material Fig. 1.**
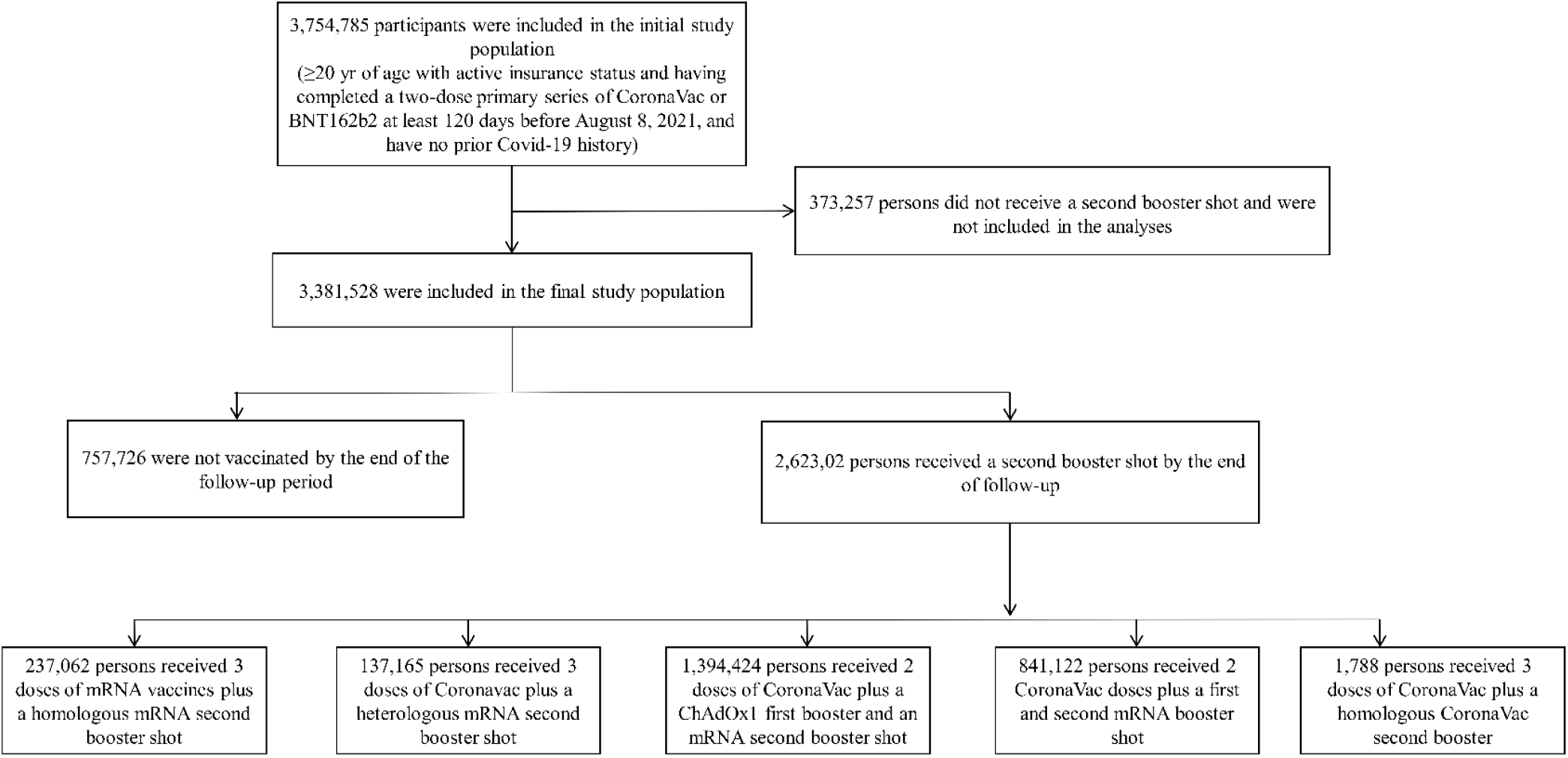
Study participants and cohort eligibility. Participants were adults aged ≥20 years affiliated with the Fondo Nacional de Salud (FONASA), the public national healthcare system in Chile, who completed a Coronavac or BNT162b2’s two-dose primary series at least 120 days before the beginning of the follow-up on August 11, 2021, when the first-booster campaign was launched and unvaccinated individuals. We excluded individuals with probable or confirmed COVID-19 according to reverse-transcription polymerase-chain-reaction assay for SARS-CoV-2 or antigen test reported before February 15, 2022. Note that 1,788 persons received three doses of CoronaVac plus a homologous CoronaVac second booster. We did not include the latter in the study due to the small sample size.

**Supplementary Material Fig. 2.**
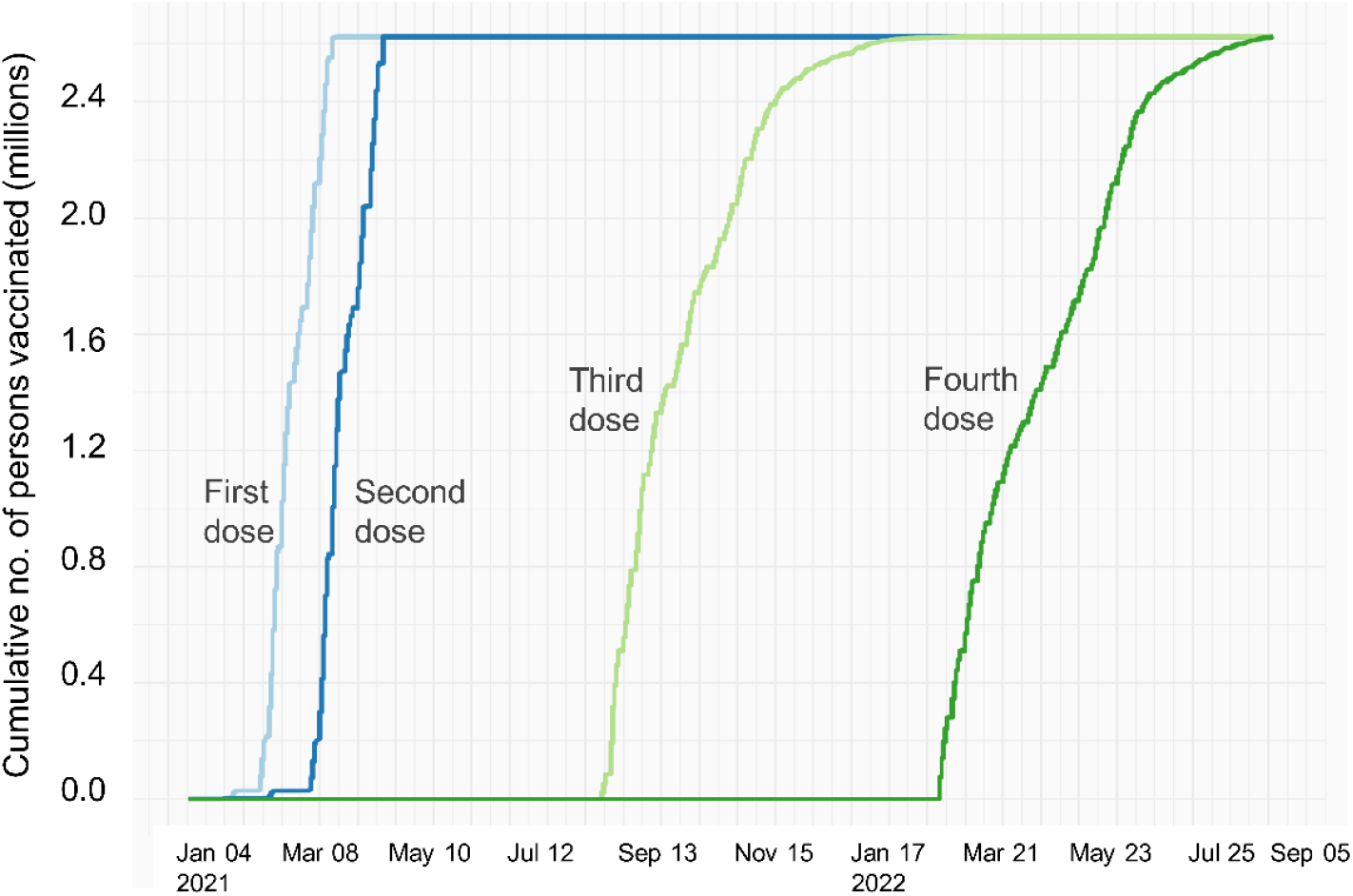
Vaccination rollout for adults aged 20 years and older who received a primary COVID-19 vaccination schedule and two booster doses. The Ministry of Health launched a COVID-19 vaccine first booster campaign on August 11, 2021, and the second booster on February 14, 2022. Our study cohort included adults aged 20 years or older affiliated with the Fondo Nacional de Salud (FONASA), Chile’s national public health insurance program.

**Supplementary Material Fig. 3.**
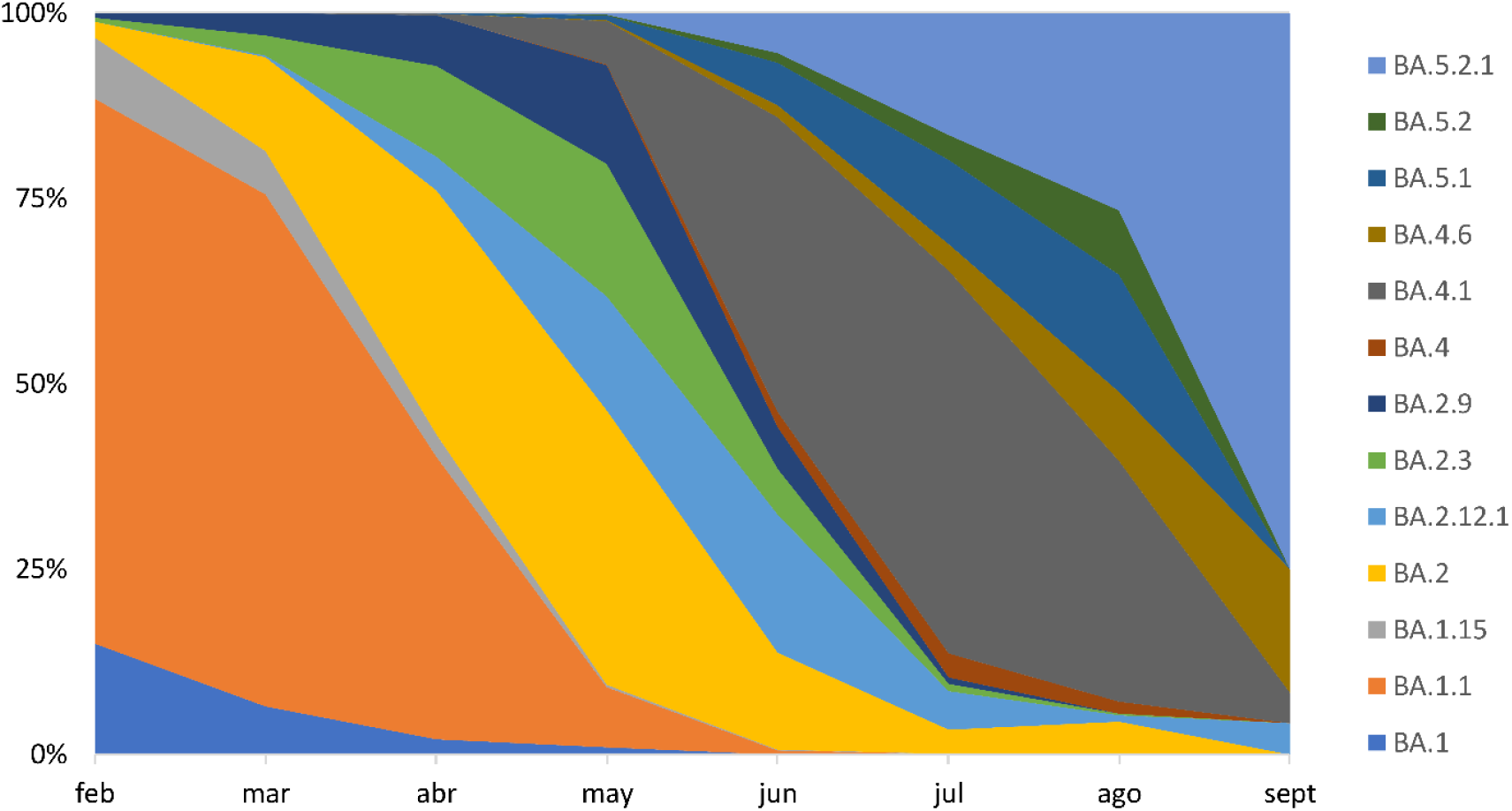
Distribution of the predominant SARS-CoV-2 lineages in Chile between February and September, 2022, based on data shared on the GISAID platform. A total of 12,578 SARS-CoV genomes were available at the GISAID EpiCov repository. The Ministry of Health monitors respiratory viruses, including SARS-CoV-2, using genomic surveillance in sentinel centers. Surveillance efforts related to the pandemic include sequencing a non-probabilistic sample of SARS-CoV-2 RT-PCR positive samples retrieved from travelers entering the country, severe COVID-19 cases, and community surveillance. During the study period, Omicron was the only variant of concern observed. The dynamics of the sub-lineages followed consecutive waves of BA.1, BA.2, BA.2.12.1, BA.4, and BA.5.

